# Correlation between Pregnancy Status and Severe Corona-Virus Disease Characterized by Cytokine Storm: Systematic Review and Meta-Analysis

**DOI:** 10.1101/2021.06.11.21258747

**Authors:** John Muthuka, Michael Kiptoo, Kelly Oluoch, Everlyn Nyamai

## Abstract

**Background:** Coronavirus disease 2019 (COVID-19) is caused by severe acute respiratory syndrome coronavirus 2 (SAR2-COV-2) that identified first in Wuhan, China, in December 2019, rapidly spreading to the rest of the globe, becoming a pandemic. Some studies have eluded to an association between pregnancy status and severe COVID-19 cytokine storm, some, in contrast, have demonstrated such. The aim of the current study was to find the relationship between pregnancy status and clinical COVID-19 severity characterized by cytokine storm through a systematic review and meta-analysis approach.

**Methods:** We searched Google Scholar, PubMed, Scopus, Web of Science, and Embase databases to identify clinical studies suitable for inclusion in this meta-analysis. Studies reporting pregnancy status and comparing the COVID-19 severity cytokine storm outcome were included. The COVID-19 severity characterized by cytokine storm was described using parameters such as; Intensive Care Unit Admission, Invasive Mechanical Ventilation, Mechanical Ventilation, Hospital Admission, Pro and Inflammatory cytokine levels, consolidation on chest CT scan, pulmonary infiltration, extreme fevers as characteristic of cytokine storm, syndromic severity, higher neutrophil count indicative of cytokine storm and severe COVI-19 presentation.

**Results:** A total of 17 articles detailing 840332 COVID-19 women were included. Our meta-analysis revealed a relationship between positive pregnancy status and severe COVID-19 cytokine storm case (random effect model, OR=2.47; 95% CI: 1.63-3.73; P < 0.0001), with a cumulative incidence of 6432 (14.1%) among the pregnant women with COVID-19 and 24352 (3.1%) among the non-pregnant women with COVI-19. Further to this, we found that the sub-analysis between Single Centre and Multiple Centre studies demonstrated seemingly the same as heterogeneity (I^2^ = 72 and (I^2^ = 98), respectively. Sensitivity analysis on each sub-group revealed that pregnancy was significantly related to severe COVID-19 with cytokine storm from single Centre studies, (fixed effect model, OR= 3.97; 95% CI: 2.26-6.95; P< 0.00001) with very low heterogeneity (I^2^ = 2 %; P = 0.42).

**Conclusion:** Being pregnant is clearly associated with experiencing a severe COVID-19 characterized by a cytokine storm. The SARS-COV-2 epidemic should serve as an impetus for pregnant women diagnosed with COVID-19, and map out salient risk factors associated with its severity. The trial is registered with the International Prospective Register of Systematic Reviews (PROSPERO) CRD42021242011.

## 1. Introduction

Coronavirus disease 2019 (COVID-19), which is caused by severe acute respiratory syndrome coronavirus 2 (SAR2-COV-2), is an infectious disease caused by a newly discovered coronavirus and was first identified in Wuhan, China, in December 2019 (Sharma, 2020). It has subsequently spread across the world, causing a global pandemic. This highly contagious disease has thus far infected over 25 million people worldwide and killed over 1 Million patients, yielding a case fatality rate (CFR) that varies between 0.7 and 12.7% (average: 3.4%) (Worldometer, 2020).

Most people infected with the COVID-19 virus will experience mild to moderate respiratory illness and recover without requiring special treatment. Older people and those with underlying medical problems like cardiovascular disease, diabetes, chronic respiratory disease, and cancer are more likely to develop serious illnesses (Handayani et al., 2020).

COVID-19 primarily targets lung epithelial cells, causing viral pneumonia and acute respiratory distress syndrome (ARDS), especially in elderly patients. Therefore, mortality is higher in the elderly and in patients with at least one accompanying comorbid disease (Yang et al., 2020). In the last report issued by the Centers for Disease Control and Prevention Institute, the incidence of respiratory disease was 9.2% in patients diagnosed with a severe COVID-19 clinical course (Chow et al., 2020). Chronic obstructive pulmonary disease (COPD) and asthma are also common comorbidities in severe cases and are reported in 10.8% and 17.0%, respectively, of hospitalized patients aged ≥18 years with COVID-19 (Garg et al., 2020).

COVID-19 infection has a heterogeneous disease course; it may be asymptomatic or causes only mild symptoms in the majority of the cases (Gerasimov et al., 2020), while immunologic complications such as macrophage activation syndrome also known as secondary hem phagocytic lymphohistiocytosis, resulting in cytokine storm syndrome and acute respiratory distress syndrome, may also occur in some patients (Hu et al., 2021). The immune system is there to help us fight infection, but sometimes it wreaks more havoc than the disease itself. Most of the COVID-19 patients experiencing cytokine storms are presenting with fevers and shortness of breath, then having so much difficulty breathing they eventually require ventilation. This event may be related to pregnancy status, as a severe presentation (Bhaskar et al., 2020)

Pregnant women who have COVID-19 appear more likely to develop respiratory complications requiring intensive care than women who aren’t pregnant, according to the Centers for Disease Control and Prevention (Ellington et al., 2020). Pregnant women are also more likely to be placed on a ventilator. Some research suggests that pregnant women with COVID-19 are also more likely to have a premature birth and cesarean delivery, and their babies are more likely to be admitted to a neonatal unit (Barile et al., 2020).

Pregnant women are a potentially highly vulnerable population due to anatomical, physiological, and immunological changes under the COVID-19 pandemic. Issues related to pregnancy with COVID-19 attracted widespread attention from researchers. A large number of articles were published aiming to elaborate on clinical characteristics and outcomes of pregnant women infected with COVID-19, in order to provide evidence for management. The existing data suggest that the overall prognosis of pregnancy with COVID-19 is promising when compared with that of other previous coronaviruses. There is still maternal morbidity and mortality related to COVID-19 reported (Chen et al., 2020).

There are many unknowns for pregnant women during the coronavirus disease 2019 (COVID-19) pandemic. Clinical experience of pregnancies complicated with infection by other coronaviruses e.g., Severe Acute Respiratory Syndrome (SARS) and Middle Eastern Respiratory Syndrome, has led to a pregnant woman being considered potentially vulnerable to severe SARS-CoV-2 infection. Physiological changes during pregnancy have a significant impact on the immune system, respiratory system, cardiovascular function, and coagulation (Wastnedge et al., 2021).

Given diverging findings in the existing literature, we systematically reviewed English-language studies to investigate whether the pregnancy was associated with a more severe clinical course of COVID-19.

## 2. Material and Methods

All guidelines listed in the Preferred Reporting Items for Systematic Reviews and Meta-Analyses (PRISMA) statement were followed in conducting this meta-analysis (Moher et al., 2016). The current methodological systematic review and meta-analysis pooled data from observational studies including cohort, case-control, cross-sectional and similar viable case studies and was recorded in the International Prospective Register of Systematic Reviews https://www.crd.york.ac.uk/PROSPERO/#myprospero; registration number: CRD42021242011

### 2.1. Study Search Strategy

We searched by a simple search in Google Scholar, PubMed, Scopus, Web of Science, and Embase databases to identify observational studies suitable for inclusion here with search terms; severe Covid-19 and pregnancy and moving on to other keywords, that is; pregnancy, severe cytokine storm, covid-19 (corona-virus, all in English for the period beginning from March 2020 to March 2021. The following search terms were used: “COVID-19,” OR “SARS-COV-2,” OR “novel coronavirus (CoV),” AND “pregnant,” OR “gestation,” AND “clinical features,” OR “characteristic,” AND “severity,” OR “severe”.

### 2.2. Inclusion and Exclusion Criteria

Inclusion criteria were as follows: (i) studies that examined COVID-19 women within reproductive age and diagnosed COVID-19 according to WHO criteria; (ii) observational, cross-sectional, prospective, or retrospective studies; (iii) studies that compared pregnant women versus non-pregnant women with severe COVID-19 characterized by cytokine storm; (iv) studies evaluating the clinical prognosis in pregnancy and the immunological profile at any gestation stage and looking at the pro-inflammatory response, covid-19 disease and the hallmark outcome, the severe cytokine storm

Exclusion criteria were as follows: (i) unrelated, duplicated, and contain information answering our research question, (ii) non-English-language studies, (iii) case reports/series, (iv) reviews, (v) editorial letters, (vi) studies lacking a full-text (unavailable or not yet published), (vii) studies without a DOI, and (viii) studies with small sample sizes (<50 patients) because of low statistical power.

### 2.3. Data Extraction

Extraction of both adjusted and non-adjusted data was be performed to give the most allowed confounding factor which would be used in the analysis by pooling them later. A researcher (JM) scanned study titles and abstracts obtained via an initial database search and included relevant ones in a secondary pool. Next, two independent researchers (MK and KO) evaluated the full text of these articles to determine whether they met study inclusion criteria. Any disputes were resolved by discussion and negotiation with a third investigator (MK). Agreed-upon studies were included in the final meta-analysis.

The following data were obtained from all studies: title, first author, publication year, location, sample size, age (median), pregnancy status (pregnant or non-pregnant), and severe COVID-19 cytokine storm presentation. Given these two groups, we were able to map out who developed the severe COVID-19 cytokine storm between them.

### 2.4. Risk of Bias (Quality) Assessment

NIH tool for observational and cross-sectional was used in this study 2–3 reviewers independently assessed the quality of the studies and added to the data extraction form before the inclusion into the analysis to reduce the risk of bias. To evaluate the risk of bias, reviewers rated each of the 14 items into dichotomous variables: yes, no, or not applicable. An overall score was calculated by adding all the items’ scores as yes equals one, while no and NA equals zero. A score was given for every paper to classify them as poor, fair, or good conducted studies, where a score from 0 to 5 was considered poor, 6 to 9 as fair, and 10 to 14 as good. Data checking was done ideally by not the ones who performed the extraction of those articles or each reviewer was assigned a different article than the one he extracted in the previous.

### 2.5. Statistical Analyses

RevMan 5.4.1 (Review Manager 5.4.1 at http://community.cochrane.org/tools/review-production-tools/revman-5/revman-5) was used to calculate odds ratios with 95% confidence intervals, which are depicted using forest plots. Quantitative numbers were measured in terms of total numbers and percentages (%). The odds ratio of severe COVID-19 cytokine storm among pregnant and non-pregnant women was calculated. Heterogeneity was evaluated with Cochran’s Q and the Higgins test. The Higgins test uses a fixed-effect model when <50%, and a random-effects model when >50%. When heterogeneity was detected, a sensitivity adjustment was made to determine its source. This procedure was performed by leaving one study out of the analysis at a time, with a fixed-effect model used after excluding heterogeneity. Subgroup, cumulative analyses, and meta-regression were used to test whether the results are consistent or not and investigate the effect of confounders on the outcome (cytokine storm) and elucidate the best predictors in pregnancy status among women with Covid-19. Publication bias was evaluated using the Cochrane Risk of Bias tool.

## 3. Results

Our initial search of international databases using the keywords described above yielded 221 articles. After excluding 70 duplicate articles, 151 articles remained. When article titles and abstracts were evaluated for appropriateness, an ultimate 29 articles met the inclusion criteria. In addition, 12 further meeting the exclusion criteria were excluded for which full texts were obtained. A total of 17 articles met all our criteria. The relevant PRISMA study flow chart is shown in Figure 1. A quality analysis of the pooled articles included in this meta-analysis was done by applying NOS. As a result, overall quality was found to be moderate (NOS score min: 5, max: 8). Results of a quality analysis of the pooled articles are available in *Table 1*.

**Figure 1.**
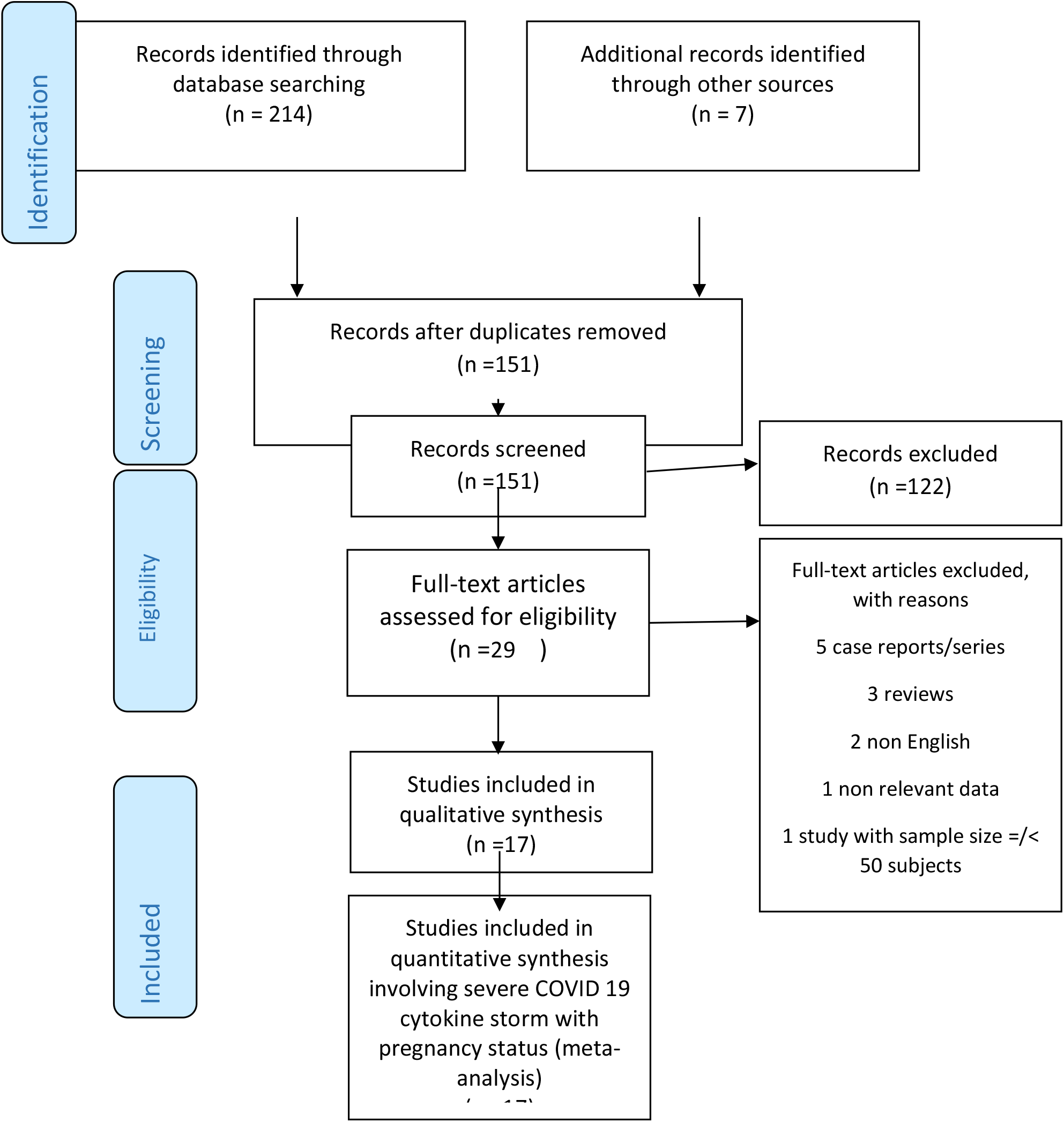
PRISMA flow diagram of study selection procedures.

### 3.1. Features of the Included Studies

In this meta-analysis pool, 840417 women diagnosed with COVID-19 from 17 studies [4, 10–24] were included. This inclusion utilized the predefined given CDC reporting guidelines on COVID-19 diagnosis (Chow et al., 2020), 85 patients whose specific parameters of consideration on the severity of COVID-19 defined with cytokine storm statuses were reported as “unknown” or not tabulated were excluded from the final analysis, yielding a final 840332 patients. Of these, 45571 (5.4%) were pregnant women diagnosed with COVID-19, while 794761 (94.6%) were non-pregnant women diagnosed with COVID-19. Among the pregnant COVID-19 diagnosed women, the total events of the severity of COVID-19 with cytokine storm were 6432 (14.1%) and among the non-pregnant COVID-19 diagnosed women were 24352 (3.1%). The total events of COVID-19 severity occurring with cytokine storm and ascertained by a specific parameter were 30784 (3.7%) with the cumulative incidence from all studies ranging from 0.4 to 90.7 % (average: 36.26%). Parameter of comparison on COVID 19 severity with cytokine storm was implicated by ICU admission in three studies, by ICU plus mechanical ventilation in two studies, three by higher levels of inflammatory response markers, one by consolidation on chest CT scan, one by pulmonary infiltration, one by extreme fevers as characteristic of cytokine storm, one by syndromic severity, one by hospital admission, two by severe COVI-19 presentation, one by invasive mechanical ventilation and one by higher neutrophil count indicative of cytokine storm. The included studies were; fifteen retrospective (six multicenter and 9 single-center) and two prospective (both multicenter). A summary of the studies included in this meta-analysis is available in Table 1.

**Table 1:** Features of the studies included in the meta-analysis.

| First author | Location of patients | Study design | Parameter of comparison on COVID 19 severity with cytokine storm | Events in Pregnant/ total in Cohort | Events in Non-Pregnant/total in cohort | Cumulative incidence of severe COVID-19 defined by cytokine storm |
| --- | --- | --- | --- | --- | --- | --- |
| Badr et al, 2020 | France and Belgium | CC, MC | ICU vs No ICU Admission | 58/83 | 17/107 | 75 [39 %] |
| Blitz et al, 2020 | New York, USA | R, O , MC | ICU vs No ICU Admission | 8/82 | 50/332 | 58 [14 %] |
| CDC, 2020 | USA | P, C, MC | ICU plus mechanical ventilation vs No ICU Admission with mechanical ventilation | 2583/8200 | 15840/316800 | 18423 [5.7 %] |
| Chen B et al 2020 | Wuhan, China | R, SC | higher level of inflammation markers vs Lower | 0/31 | 1/80 | 1 [0.9 %] |
| Collin et al, 2020 | Sweden | R, MC | invasive mechanical ventilation vs no invasive mechanical ventilation | 7/13 | 29/40 | 36 [68 %] |
| ELLINGTON et al, 2020 | USA | R, O MC | ICU with MV vs no ICU MV | 2587/8207 | 4840/83205 | 7427 [8 %] |
| Fang Liu et al, 2020 | Wuhan, China | R, CC, SC | Consolidation on chest CT vs no consolidation on chest CT | 20/21 | 16/19 | 36 [90 %] |
| MartinezPortilla et al 2020 | Mexico | R, MC | ICU/Death vs non ICU/Death | 752/5183 | 446/5183 | 1198 [12 %] |
| Ming-Zhu Yin et al, 2020 | China | R,C,SC | Severe or critical COVID-19 characterized by higher levels of inflammatory indices of cytokine storm vs Moderate COVID-19 | 19/31 | 11/35 | 30 [46 %] |
| Mohr-Sasson et al, 2020 | Fuyang, China | R, C, SC | High vs low fevers | 3/11 | 15/25 | 18 [50 %] |
| Molteni E, et al 2020 | UK, SWEDEN & USA | P,O , MC | syndromic severity vs non syndromic severity | 87/140 | 1508/2515 | 1595 [60 %] |
| Oakes et al, 2020 | Wuhan, China | R, C, SC | Hospital admission vs non-admission | 7/22 | 17/240 | 24 [9 %] |
| Qiancheng et al, 2021 | Wuhan, China | R, SC | Non severe vs. severe | 2/28 | 1/54 | 3 [9.8 %] |
| Wang Z et al 2020 | Wuhan, China | R, SC | Covid-19 Manifestations on chest CT/ non | 22/30 | 42/42 | 64 [89 %] |
| Wei L et al 2020 | Wuhan, China | R, SC | Higher vs lower neutrophil count as indicative of cytokine storm | 15/17 | 24/26 | 39 [91%] |
| Xu S et al 2020 | Wuhan, China | R, , SC | Pulmonary infiltration vs no pulmonary infiltration | 17/34 | 3/30 | 20 [31 %] |
| Zambrano et al, 2020 | USA | R, MC | Severe COVID-19-associated illness vs Mild to moderate | 245/23434 | 1492/386,028 | 1737 [0.4 %] |

*R: retrospective; CS: cross-sectional; O: observational; ICU: intensive care unit; SC: single-center; MC: multicenter; CC: case-control; P: prospective;*

The main outcome of this meta-analysis was the possible association of pregnancy and severe COVID-19 characterized by cytokine storm which was indicated by a specific prognosis and event such as, ICU admission or such as shown in Table 1. We assessed the quality of the included observational studies based on a modified version of the Newcastle-Ottawa Scale (NOS) which consists of 8 items with 3 subscales, and the total maximum score of these 3 subsets is 9. We considered a study that scored ≥7 a high-quality study since a standard criterion for what constitutes a high-quality study has not yet been universally established. The 17 studies assessed by us generated a mean value of 6.47 and as result, the overall quality was found to be moderate (NOS score min: 5, max: 8).as indicated in Table 2.

**Table 2:** Newcastle-Ottawa scale for quality assessment and risk of bias.

| Study/Year | Year | Case selection (max. 4) | Comparability (max. 2) | Exposure/outcome (max. 3) | Total score |
| --- | --- | --- | --- | --- | --- |
| (Badr et al., 2020) | 2020 | *** | ** | ** | 7 |
| (Blitz et al., 2020) | 2020 | *** | ** | * | 6 |
| (Centers for Disease Control and Prevention, 2020) | 2020 | **** | ** | ** | 8 |
| (Cheng et al., 2020) | 2020 | *** | * | ** | 6 |
| (Collin et al., 2020) | 2020 | **** | * | ** | 7 |
| (Ellington et al., 2020) | 2020 | *** | ** | *** | 7 |
| (Liu et al., 2020) | 2020 | *** | * | ** | 6 |
| (Martinez-Portilla et al., 2021) | 2020 | *** | * | ** | 6 |
| (Yin et al., 2020) | 2020 | *** | ** | ** | 7 |
| (Mohr-Sasson et al., 2020) | 2020 | *** | * | * | 5 |
| (Molteni et al., 2020) | 2020 | *** | * | ** | 6 |
| (Oakes et al., 2021) | 2020 | *** | ** | ** | 7 |
| (Qiancheng et al., 2020) | 2021 | *** | * | ** | 6 |
| (Wang et al., 2020) | 2020 | ** | ** | ** | 6 |
| (Wei et al., 2020) | 2020 | *** | * | *** | 7 |
| (Xu et al., 2020) | 2020 | *** | ** | ** | 6 |
| (Zambrano et al, 2020) | 2020 | *** | * | *** | 7 |

### 3.2. Pregnancy status and COVID-19 Severity characterized by Cytokine Storm

The cumulative incidence of severe COVID-19 characterized by Cytokine Storm as ascertained by a specific indicator among pregnant women was 6432/45571(14.1%), while among non-pregnant women, it was 24352/794761 (3.1%). A meta-analysis revealed a significant association between pregnancy status and severe COVID-19 with cytokine storm (random effect model, OR=2.47; 95% CI: 1.63-3.73; P < 0.0001) (Table 3) (Figure 2(a)). A funnel plot was used to evaluate publication bias and revealed considerable heterogeneity between all the pooled studies (I^2^ = 98 %; P < 0.00001) (Figure 2(b)). A sensitivity analysis was performed to explore the impact of excluding or including the cumulative incidence of severe COVID-19 characterized by Cytokine Storm as ascertained by a specific indicator among pregnant women was 6432/45571(14.1%), while among non-pregnant women, it was 24352/794761 (3.1%). A meta-analysis revealed a significant association between pregnancy status and severe COVID-19 with cytokine storm (random effect model, OR=2.47; 95% CI: 1.63-3.73; P < 0.0001) (Table 3) (Figure 2(a)). A funnel plot was used to evaluate publication bias and revealed considerable heterogeneity between all the pooled studies (I^2^ = 98 %; P < 0.00001) (Figure 2(b)). A sensitivity analysis was performed to explore the impact of excluding or including studies in a meta-analysis based on sample size, methodological quality, and variance. After removing eight studies (Blitz et al., 2020; Centers for Disease Control and Prevention, 2020; Collin et al., 2020; Martinez-Portilla et al., 2021; Mohr-Sasson et al., 2020; Molteni et al., 2020; Wang et al., 2020; Zambrano et al., 2020), which were major causes of heterogeneity, revealed that pregnancy was significantly related to severe COVID-19 with cytokine storm (fixed effect model, OR= 7.41; 95% CI: 7.02-7.83; P< 0.00001). Furthermore, this updated analysis showed substantially low heterogeneity (I^2^ = 29 %; P = 0.19;), while a funnel plot revealed no publication bias (Figure 2(d)).g studies in a meta-analysis based on sample size, methodological quality, and variance. After removing eight studies (Blitz et al., 2020; Centers for Disease Control and Prevention, 2020; Collin et al., 2020; Martinez-Portilla et al., 2021; Mohr-Sasson et al., 2020; Molteni et al., 2020; Wang et al., 2020; Zambrano et al., 2020), which were major causes of heterogeneity, revealed that pregnancy was significantly related to severe COVID-19 with cytokine storm (fixed effect model, OR= 7.41; 95% CI: 7.02-7.83; P< 0.00001). Furthermore, this updated analysis showed substantially low heterogeneity (I^2^ = 29 %; P = 0.19;), while a funnel plot revealed no publication bias (Figure 2(d)).

**Table 3:**
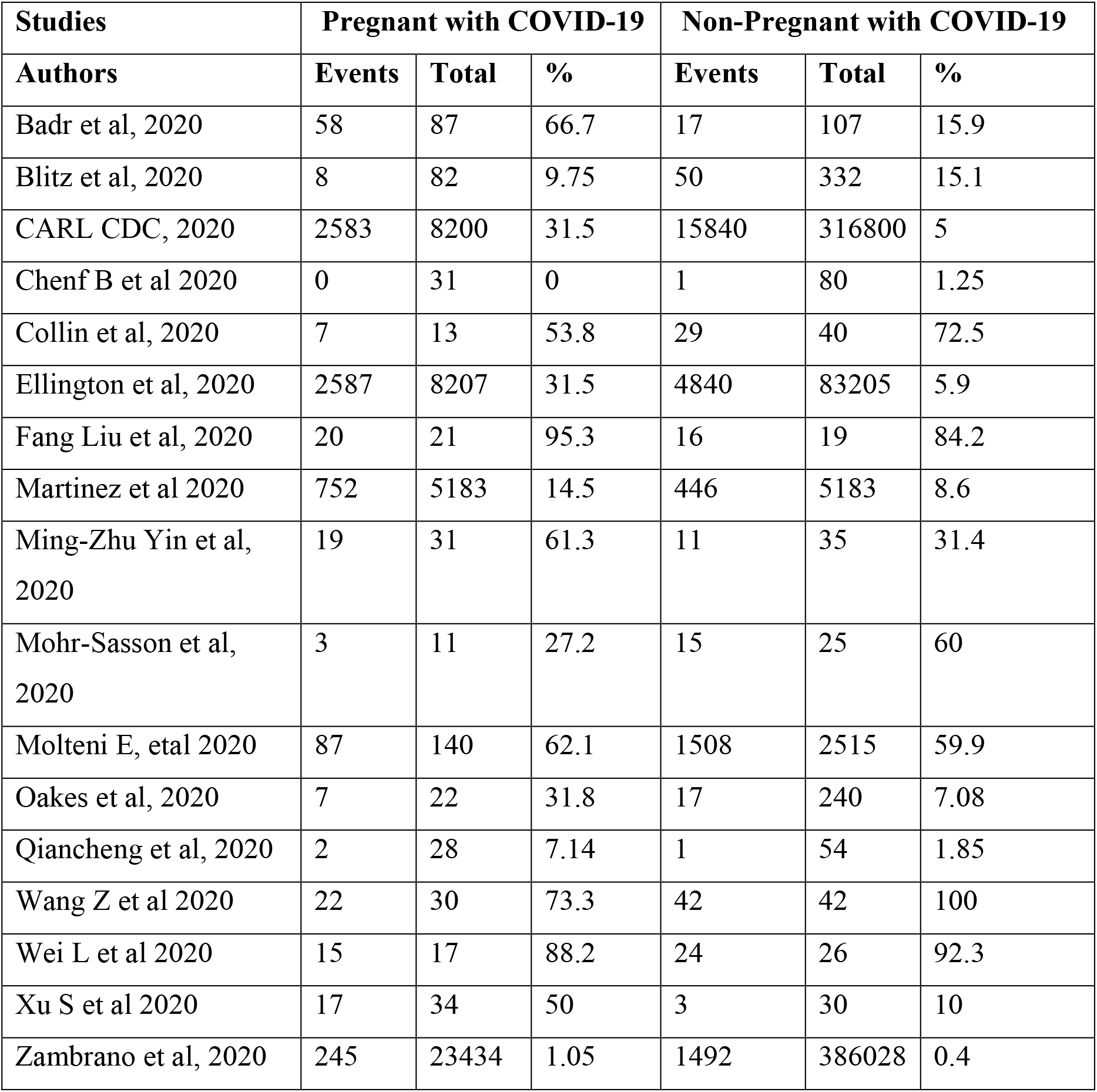
Events in pregnant and non-pregnant women.

**Figure 2.**
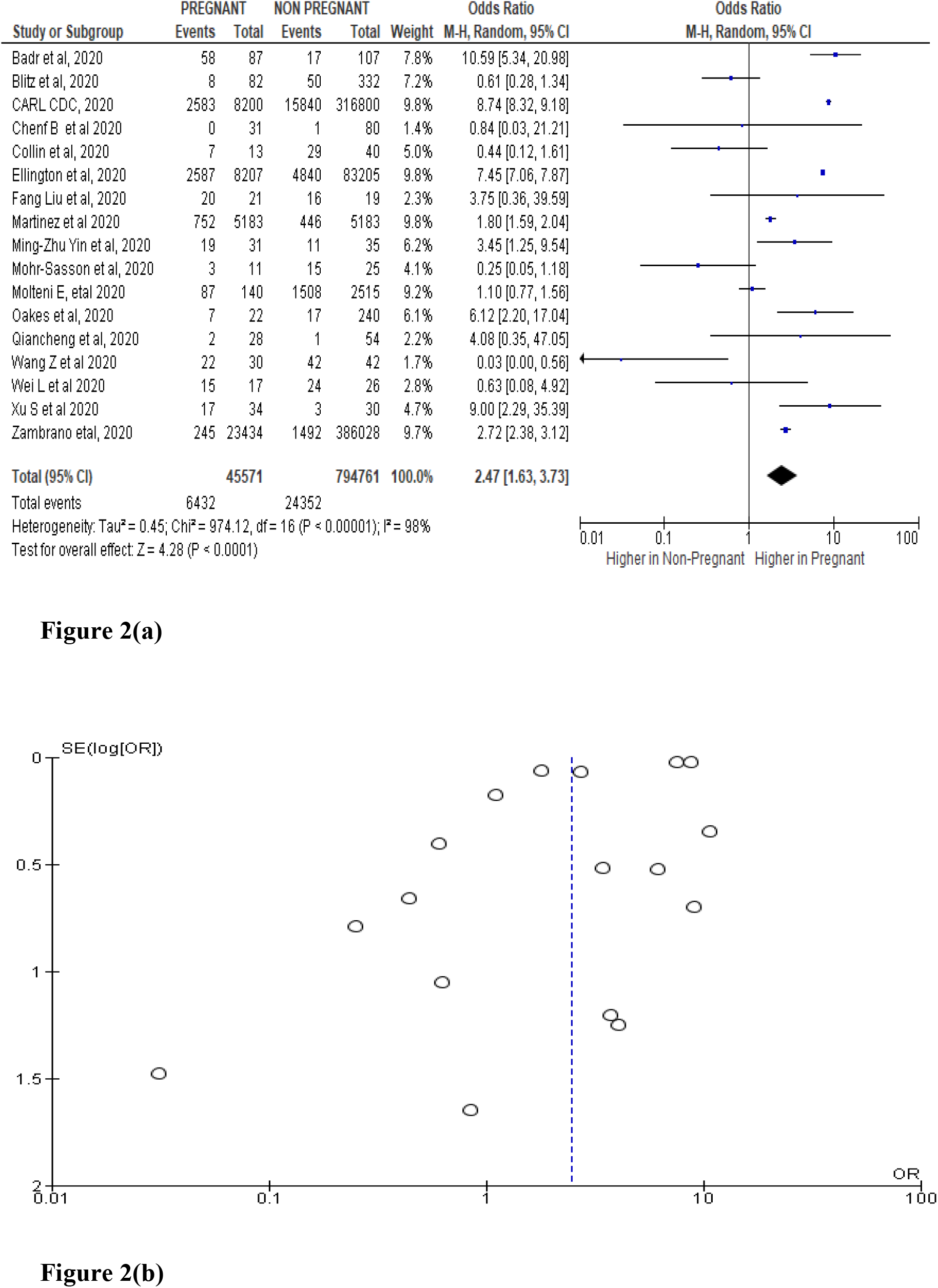
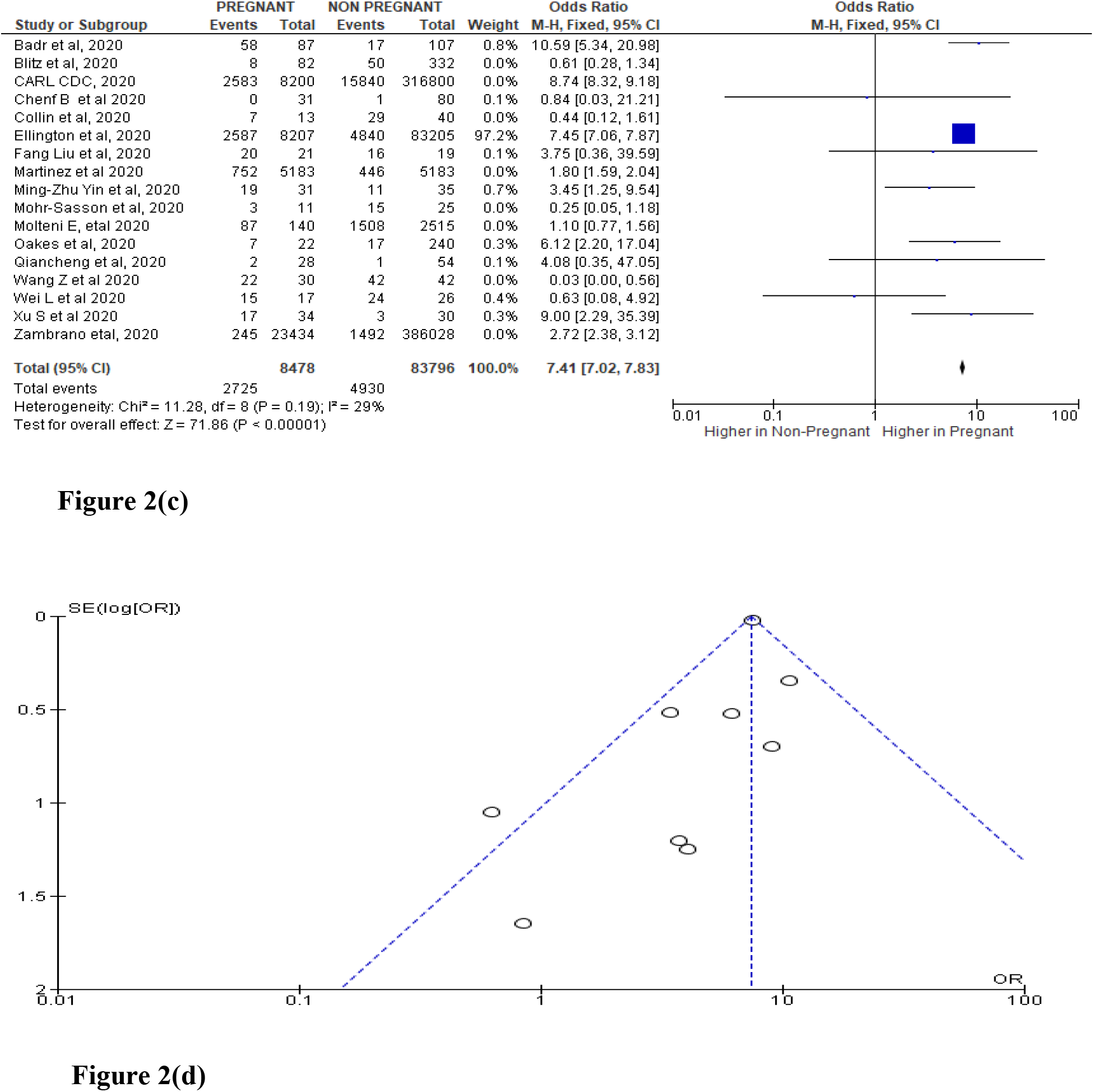
(a) The cumulative incidence of severe COVID-19 characterized by Cytokine Storm. (b) Funnel plot for meta-analysis of cumulative incidence of severe COVID-19 characterized by Cytokine Storm. (c) The cumulative incidence of severe COVID-19 characterized by Cytokine Storm after excluding or including studies in meta-analysis based on sample size, methodological quality and variance. (d) Funnel plot for the meta-analysis of excluding or including studies in meta-analysis based on sample size, methodological quality and variance.

### 3.3 Subgroup analysis and investigation of heterogeneity

Heterogeneity in the pooled effect estimates was considerably high in all the 17 studies and thus, it was necessary to perform subgroup analyses to identify possible variables or characteristics moderating the results obtained. This was done based on the number of Centers that the data was obtained from, as either Multiple Centre (MC) or single Centre studies (SC). The Subgroup Analysis demonstrated that, they were seemingly the same as heterogeneity was still quite high (I2 = 72), Test for overall effect: Z = 0.91 (P = 0.36) and (I2 = 98), Test for overall effect: Z = 3.97 (P < 0.0001) for SC and MC respectively both with P<0.00001 with no significance difference (test for subgroup differences: Chi^2^ = 0.67, df = 1 (P = 0.41), I^2^ = 0%) (Figures 3(a) and 3(b). This prompted further sensitivity analysis on each subgroup to ascertain the group that mostly was associated with heterogeneity.

**Figure 3.**
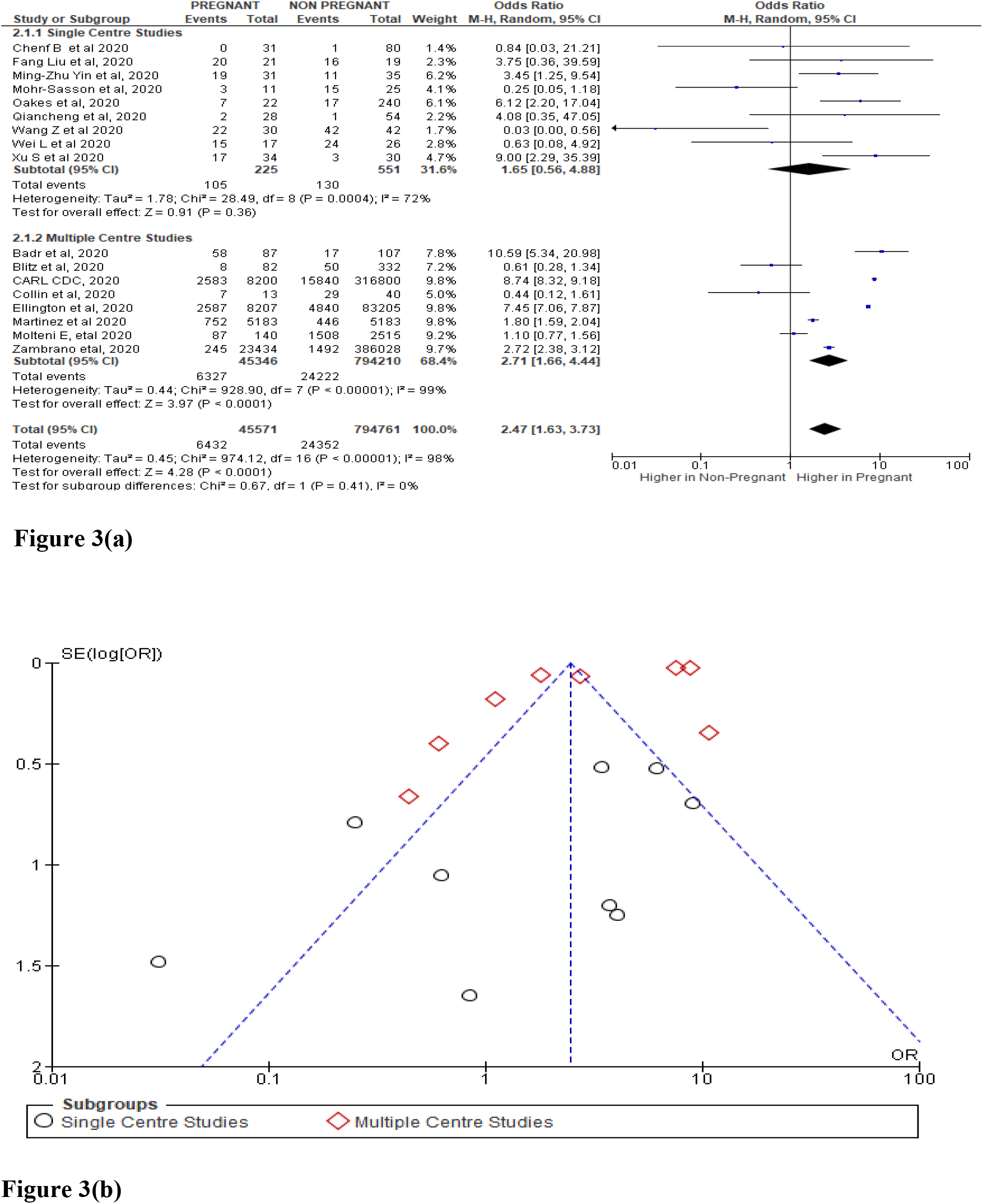
(a) Subgroup analysis between Single Centre and Multiple Centre studies (b) Funnel plot for the meta-analysis of pregnancy and severe COVID-19 disease characterized by cytokine

The sensitivity analysis on independent sub-analysis groups showed that Single Centre studies with the elimination of studies that were causing the major heterogeneity (Mohr-Sasson et al. [] and Wang Z et al. [], revealed that pregnancy was significantly related to severe COVID-19 with cytokine storm from single Centre studies, (fixed effect model, OR= 3.97; 95% CI: 2.26-6.95; P< 0.00001) with this updated analysis showing substantially very low heterogeneity (I^2^ = 2 %; P = 0.42).

In regards to the Multiple Centre studies, subsequent removal of any study did not change the heterogeneity from its original (Heterogeneity: Chi^2^ = 928.90, df = 7 (P < 0.00001); I^2^ = 99%), demonstrating that, Multiple Center studies were the main cause of heterogeneity and this was similar to the overall heterogeneity of the combined groups (Fixed effects model, Heterogeneity: Chi^2^ = 938.26, df = 14 (P < 0.00001); I^2^ = 99%), with the test for subgroup differences being: Chi^2^ = 1.9, df = 1 (P = 0.17), I^2^ = 47.4%) (Figure 4(a)). The funnel plot similarly demonstrated that, MC studies were associated with heterogeneity with only one study forming part of homogeneity (Figure 4(a).

**Figure 4.**
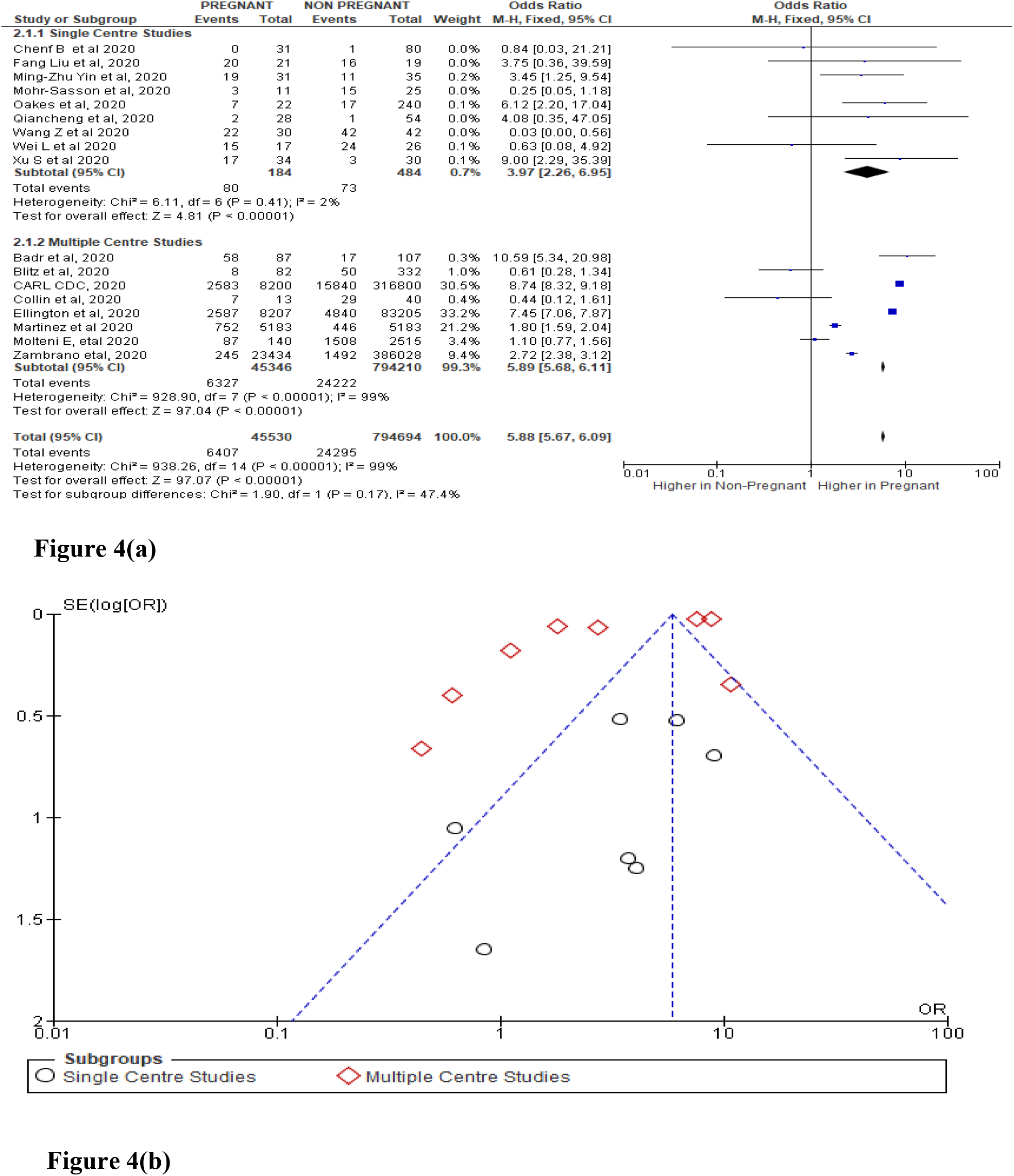
(a) Comparison of specific subgroup analysis sensitivity testing between Single Centre and Multiple Centre studies in severe COVID-19 patients. (b) Funnel plot for the meta-analysis of pregnancy with severe COVID-19 disease between the subgroup analysis (Single Centre and Multiple Centre studies)

## 4. Discussion

The present meta-analysis contained 17 studies and revealed that pregnant women had significantly increased risk for severe COVID-19, characterized by a cytokine storm. Previous research has seemingly reported a similar result (Czeresnia et al., 2020; Kolkova et al., 2020). Additionally, one meta-analysis study has reported the outcome of coronavirus spectrum infections (SARS, MERS, and COVID-19) during pregnancy that, COVID-19 disease severity increased in gestation (Di Mascio et al., 2020). This analysis adds to the extensive consensus in the literature, invoking more studies and examining pregnancy status as a possible predictor of severe COVID-19 characterized by a cytokine storm.

Prior studies have reported results that contrast with those presented here—namely, no significant difference between pregnant versus non-pregnant women diagnosed with COVID-19 in terms of its associated severity (Selim et al., 2020; Shankar & KR, 2020; TIRMIKÇIOĞLU, 2021; TN & RA, 2020). In addition, a meta-analysis conducted by Matar et al. (Matar et al., 2021) failed to find a relationship between being pregnant and severe COVID-19 24 studies including pregnant women, and another meta-analysis indicates that COVID-19 infection during pregnancy probably has a clinical presentation and severity resembling that in non-pregnant adults (Elshafeey et al., 2020). Also, a meta-analysis compared similar trends in severity between pregnant and the general population (Kasraeian et al., 2020). Further, two more studies showed no feasible differences in the clinical presentation of COVID-19 between pregnant and non-pregnant women (Jafari et al., 2021; Vaezi et al., 2021). Of concern, both meta-analyses (Elshafeey et al., 2020; Matar et al., 2021) included no assessment of publication bias or study quality. As such, it should be considered only a preliminary quest. Hence, the present systematic meta-analysis offers a more detailed view as it covers 17 studies from diverse regions capturing both single and multiple centers. The heterogeneity was high, and after sensitivity adjustments, the incidence of COVID – 19 severity with pregnancy was imminent with a substantially low heterogeneity after eliminating studies causing the same. Furthermore, the sub-group analysis after performing the sensitivity test in each specified subgroup (either MC or SC), there was a clear significant association between being pregnant and developing severe COVID-19 characterized by any specific parameter of cytokine storm evidenced in SC studies. Therefore, severe COVID-19 was observed almost 4 times (OR= 3.97; 95% CI: 2.26-6.95; P< 0.00001) more frequently in pregnant women. Some conducted studies including some meta-analysis (Al-Matary et al., 2021; Galang et al., 2020; Grünebaum et al., 2021; Kasraeian et al., 2020; Lucarelli et al., 2020; Soheili et al., 2021; 2020, 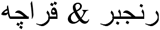) results have seemingly supported the current findings.

A recent meta-analysis revealed that SARSC-CoV-2 infection may not behave as mild as suggested during pregnancy (Barbero et al., 2020). Interestingly, this meta-analysis indicated that showed that 40 patients developed pneumonia, bilateral in most cases, with a 46.2% rate of hospitalization and 4 patients requiring intensive care unit (ICU) admission. The study found a higher rate of coronavirus disease (COVID-19) severe forms, even when compared to non-pregnant women with the same baseline characteristics (Barbero et al., 2020). This appears to be because pregnant women during the gestation period encounter a form of pro-inflammatory and inflammatory episodes which seem to mimic the trends of cytokine storm in Sever COVID-19 disease. This has been demonstrated in recent past studies (Cavalcante et al., 2021; Da Silva et al., 2020; Figuero et al., 2020; Malinowski et al., 2020). In addition, pregnancy has been reported to increase the progression of COVID-19 disease (Dashraath et al., 2020). There is growing evidence to support WHO’s statements that pregnant women are at a higher risk of developing severe COVID-19 related symptoms and possible mortality (Blauvelt et al., 2020; Oganyan et al., 2020; Rashid et al., 2020; San-Juan et al., 2020). Again, pregnancy has been found to worsen the morbidity of COVID-19 and this effect becomes more prominent as pregnancy advances (Tug et al., 2020).

As it is of now, its somehow clear that postponing pregnancy during the pandemic period is imperative due to the possibility of encountering worse clinical outcomes and complications associated with pro-inflammatory and inflammatory cytokines creating synergy for severe COVID-19 cytokine storm(Ghi et al., 2020; Malinowski et al., 2020; Naidu et al., 2020).

The association between pregnancy and illness severity due to other respiratory viruses has been investigated previously. In one study, in pregnancy, the case fatality rate of 25%, ICU admission (50%), and mechanical ventilation (33%), compared with the non-pregnant population (20%) was experienced(Martens & Kalgi, 2016), which may be implicated by immune responses in pregnancy. In addition, pregnancy may propagate respiratory infections and increase the risk of hospitalization (Lokken et al., 2021). A study also eluded that, complications of severity with other SARD are enhanced in pregnancy (Wastnedge et al., 2021). As a result, adverse effects on the pregnant woman’s lungs may aggravate the symptom severity of viral infections.

The novel SARS-CoV-2 uses an angiotensin-converting enzyme (ACE)-2 in the lungs to enter cells and cause infection. ACE2 expression and activity are enhanced during pregnancy, the transient ACE2 overexpression and increased activity during pregnancy may be important in modulating systemic, as well as local hemodynamics in the uteroplacental unit (Brosnihan et al., 2004; Levy et al., 2008). The ACE-2 upregulation may increase infectiousness and therefore infection severity risk, as the SARS-COV-2 virus uses this receptor for host entry. Paradoxically, it is stated that it may be useful in protecting people from acute lung injury (Pathangey et al., 2021).

In one recent study, ACE-2 gene expression was upregulated and widespread in specific (Li et al., 2020), thereby suggesting a mechanism by which risk for severe COVID-19 increases in pregnancy. The role of ACE2 in COVID-19 pathophysiology, including factors influencing ACE2 expression and activity in relation to COVID-19 severity, has been evidenced (Bourgonje et al., 2020), and thus, should be investigated for their potential impact on ACE-2 expression and thus SARS-COV-2 entry into the host and such, much focus in pregnancy (South et al., 2020).

Cytokine storm has received more attention because of the COVID-19 pandemic. Although we are learning more every day, cytokine storm seems to be at least part of the reason some people develop life-threatening symptoms from COVID-19, the medical condition caused by infection with SARS-CoV-2, and also, Hyper inflammatory cytokine storms in many severely symptomatic Covid-19 patients may be rooted in an atypical response to SARS-CoV-2 by the dysfunctional MCs of MCAS rather than a normal response by normal MCs (Afrin et al., 2020). This may be explained by systemic and chronic inflammation, diminished respiratory function and capacity, and COPD-related respiratory failure in some patients. There have been findings stipulating the association of pro-and anti-inflammatory cytokines which play crucial roles in the development and function of preeclampsia (Aggarwal et al., 2019). Given this, pregnancy itself and pro and inflammatory cytokines should be considered together as a single risk factor for severe COVID-19 among pregnant women diagnosed with the novel coronavirus.

Another critical area of concern is that cytokine storms, which are seen in some severe COVID-19 patients, are key causes of mortality in COVID-19. In these patients, pro-inflammatory cytokines such as interleukin (IL)-1, IL-2, IL-6, IL-8, IL-17, interferon-γ, and TNF-α are elevated, which affect the patient’s clinical symptoms and severity in the general population (McGonagle et al., 2020). In pregnancy, immune responses that are necessary to promote healthy pregnancy and those that lead to congenital disorders and pregnancy complications, with a particular emphasis on the role of interferons and cytokines are obvious (Yockey & Iwasaki, 2018), some being similar to the ones activated during COVID-19 cytokine storms such as TNF-α, IL-1β, and IL-6 which are some of the fundamental cytokines that function in blastocyst implantation (Al Jameil et al., 2018). Therefore, increased levels of INF-γ, LH, and prolactin having been identified as the underlying cause for recurrent pregnancy losses, therefore not only the severity amplification of cytokine storm of COVID-19 in pregnant women but consequential adverse pregnancy outcomes (Al Jameil et al., 2018). This potential interaction should be clarified with future clinical research, all the same.

Several factors limit the interpretation of the present study. First, the vast majority of studies included here were retrospective epidemiological studies conducted in the majorly USA and China, although with others from some different regions. Second, some included studies did not distinguish the age range of the participants as well as the stage of the gestation period. Third, CCVID 19 severity as assumed to be characterized by cytokine storm relied on different parameters of clinical implications such as; level of inflammatory cytokines, invasive mechanical ventilation, ICU admission, and such. Given these limitations, caution should be exercised while interpreting the current findings for more valid clinical practice. Future studies may respond to these issues by defining disease severity more clearly and by obtaining more detailed information on the associated inflammatory cytokines defining the COVID-19 cytokine storm.

Factors responsible for recurrent pregnancy loss are multiple and altered cytokine profile results in loss of pregnancy especially in the early stages of gestation. Similarly, exposure to high maternal pro-inflammatory cytokine concentrations in early pregnancy might play a part in several futuristic adverse effects on either the woman or the infant outcomes. With future studies, there is a need of the hour that women expecting a pregnancy must be screened to assess the cytokine profile even before conception to avoid loss of pregnancy and to improve the health and social well-being of the females as this may be aggravated in COVID-19 severity.

Finally, the interactions between the inherent inflammatory cytokines and cytokine storm due to COVID 19 should also be examined and clarified. In addition, clinicians can pay more attention to the history of pregnancy-related altered immune responses of COVID-19 patients, and more further research may aim to determine mechanisms that drive or decrease this risk of the severity by a ‘within’ the pregnant population study approach.

## Conclusion

The present meta-analysis revealed that pregnancy is significantly associated with increased COVID-19 symptom severity defined by a cytokine storm. The SARS-COV-2 epidemic should serve as an impetus for pregnant women diagnosed with COVID-19, and map out salient risk factors associated with its severity with an aim of maintaining good health pregnancy outcome and possibly if at all, evade COVID-19 adverse clinical prognosis.

## Data Availability

The data used to support the findings of this study are available from the corresponding author upon request.

## Conflicts of Interest

The authors have stated explicitly that there are no conflicts of interest in connection with this article.

## Authors’ Contributions

JM performed the concept development of the manuscript, data collection, collation and retrieval, design of tables, images, and figures, major analysis and interpretation of data and reporting, and writing and drafting of the manuscript; KO performed the data collection and quality assessment; EN performed the acquisition of data, quality analysis, writing and revision of the manuscript; MK performed the major role in reviewing and revising the manuscript.

## Acknowledgments

Funding of the Kenya Medical Training College for the execution of this study is gratefully acknowledged.

## Notes

### Competing Interest Statement

The authors have declared no competing interest.

